# Barriers and facilitators to successful transition to civilian life for ex-servicewomen: the perspective of service providers and policymakers

**DOI:** 10.1101/2024.08.30.24312844

**Authors:** Bethany Croak, Laura Rafferty, Marie-Louise Sharp, Alexandria Smith, Rafiyah Khan, Victoria Langston, Neil Greenberg, Nicola T Fear, Sharon A.M Stevelink

**Affiliations:** King’s Centre for Military Health Research, King’s College London, London, UK; Department of Psychological Medicine, Institute of Psychiatry, Psychology and Neuroscience, King’s College London, London, UK; Academic Department of Military Mental Health, King’s College London, London, UK

**Author notes:** Corresponding author: Bethany Croak. King’s Centre for Military Health Research, King’s College London, Weston Education Centre, Cutcombe Road, London SE5 9RJ.

## Abstract

The role of women in the UK Armed Forces has changed considerably in the last decade. With drives to increase the number of women serving in the military, research must consider the impact of both service and transition into civilian life on the health and wellbeing of service and ex-servicewomen (female veterans). This paper adds to the field by providing the perspective of service providers supporting ex-service personnel with their mental health, employment, housing and other needs in addition to those working in policy affecting ex-servicewomen. This study aimed to explore their understanding of what constitutes a successful transition into civilian life, the barriers and facilitators to achieving this and how transition might be impacted by the gender of the individual transitioning. Interviews and roundtable discussions were held with stakeholders (n=28) and analysed using framework analysis. Four overarching themes were identified: ’Successful transition is individual and all-encompassing’, ‘The conflicting identities of servicewomen’, ’Sexism: women don’t belong in service’ and ’The needs of servicewomen’. The first theme describes how the process and result of successful transition is individual to each ex-servicewoman, whilst the remaining themes outline common challenges faced by ex-servicewomen on this journey. There was no singular definition of ’successful transition’, but stakeholders described barriers to a successful transition. They identified prominent gender-specific barriers rooted in misogyny and inequality during military service that permeated into civilian life and impacted support use and workplace experiences. Ex- servicewomen were often required to juggle multiple responsibilities, mother and partner, and identities, women and warrior, simultaneously. Policies should look to address elements of military culture that may reinforce gender inequality and ensure veteran services are inclusive and welcoming to women and cater for gender-specific needs such as gynaecological health. Whilst in-service and veteran-focused interventions are needed, entrenched sexism in general society should not be ignored.

## 1. Introduction

Women make up approximately 11% of the UK Armed Forces (1) and the UK Government aims to increase this to 30% by 2030 (2). The role of women in the UK Armed Forces has changed substantially since the inception of The Women’s Army Auxiliary Corps in 1917 through to 2016, when women were first allowed to join close ground combat roles (3). Whilst both the role and extent of women in the Armed Forces has changed, the research evidence pertaining to women in the Armed Forces has not kept pace. The concentration of research centred on male service members has left a knowledge gap about what happens to servicewomen when they leave the Armed Forces, and there is a lack of understanding of how a career in the military, a male-dominated environment, impacts women.

The majority of ex-service personnel^1^ (veterans) transition to civilian life well. The vast majority (87%) of ex-service personnel are employed six months after leaving service (4) and although ex-servicewomen are less likely than their male counterparts to be employed (78%), this reflects employment rate differences seen in the general population (4). Another popular indicator of transition outcomes is housing; personnel may be living in military accommodation when they leave and so finding a home outside the military is essential. Data from the UK census in 2021 indicates that 1.88% of residents at homeless hostels or shelters had served in the UK Armed Forces; this is lower than the proportion of the population of England and Wales who are veterans (3.81%) (5). In addition to practical needs, health after service is often scrutinised when looking at transition. Leaving the military and transitioning into civilian life can be a substantial emotional adjustment for some service personnel. Ex-service personnel report experiencing confusion with their own identity, a feeling of loss, and a disconnect from society (6). Lower levels of social integration in ex-service personnel have been associated with common mental disorders (CMD) such as anxiety and depression (7). Extant evidence suggests that women (service and ex-service) have similar rates of Post-Traumatic Stress Disorder (PTSD) as service and ex-servicemen (8).

On the surface, general outcomes seem to appear similar for men and women after service. However, the dearth of research into women’s experiences specifically means that subtle complexities might be overlooked. For example, whilst the rates of PTSD are similar amongst service and ex-servicemen and women, it is possible that there may be differences in the type of trauma they’re exposed to. A meta-analysis from the United States (US) found that 38.4% of service and ex-servicewomen reported Military Sexual Trauma (MST) comprised of harassment and assault, compared to 3.9% of service and ex-servicemen. For sexual assault alone, the reported prevalence was 23.6% for service and ex-servicewomen assault compared to 1.9% for service and ex-servicemen (9). Although there is no evidence to show the prevalence of MST amongst UK service or ex-service population as a whole, evidence from a sample of help-seeking ex-servicewomen, showed a high prevalence of in-service sexual harassment (22.5%), sexual assault (5.1%), emotional bullying (22.7%) and physical assault (3.3.%) (10). Those who experienced these adversities had a greater risk of PTSD (10). If MST is as pervasive in the whole UK service population as it is in the US, servicewomen are at risk of poor mental health outcomes that may require significant support and treatment after they leave the military and rejoin civilian life.

In addition to potential differences in in-service experience across genders, it is possible that there are gender-specific factors that impact the experience of transition, yet this is not well understood. One study that explored transition challenges beyond service found that women perceived it to be harder to develop a post-service social network and transition to civilian life than men, and that they felt more uncomfortable attending military veteran events than their male counterparts (11). However, no research to our knowledge has verified this perception.

In light of the increasing role women play in the Armed Forces, and the potentially different experiences that they may face, it is imperative that the service provision for veterans supports both men and women. This involves supporting both different mental health experiences and differences in mental health help-seeking. In general, ex-servicewomen do seek help for their mental health and are, in fact, more likely to seek formal support than ex-servicemen, yet they still face barriers to mental health help-seeking. Ex-servicewomen report similar levels of structural and attitudinal help-seeking barriers as men; in particular, self-stigma (12).

Conversely, whilst a desire not to be seen as weak is a barrier reported by both men and women, a study found that women particularly felt this pressure because of sexism faced by male colleagues whilst in service who believed women were weak (13). Although women do seek help, this might help to explain why ex-servicewomen are more likely to use generic civilian services as opposed to veteran-specific services (13). Research has further found that veteran-specific services are perceived to be male-dominated, designed for men and unaware of women’s needs (14, 15).

Ensuring that ex-servicewomen face no disadvantage when leaving service is a priority for Government and policymakers not only because of a duty of care to those who serve to protect the nation, as enshrined in the Armed Forces Covenant (16) but also because positive outcomes of military service may make it an attractive career to potential female recruits, necessary to meet the Government’s 30% target (2). The research landscape regarding transition tends to focus on the key indicators of successful transition: employment, housing and health. However, no research to our knowledge has investigated whether women have different priorities or what their perspective is on what it means to transition well when they leave the Armed Forces. Without such an understanding, it is difficult for service providers and policymakers to know their intended aims and outcomes when designing services and policies. Additionally, they are ill informed to determine which outcomes are most appropriate to measure to understand their effectiveness in supporting a successful transition.

This research forms part of a wider project that seeks to understand what is meant by a successful transition from the UK Armed Forces for ex-servicewomen, how the transition experience may be impacted by gender, and identify barriers and facilitators to a successful transition for UK ex-servicewomen. This paper reports on the veteran service provider perspective. Support service providers and those who work in relevant policy areas and clinical service delivery in Government, charitable and statutory bodies (called stakeholders from this point onwards) can offer valuable insight as rather than discussing one individual experience, they can reflect on the various ex-servicewomen they have supported, helping to identify patterns and commonalities regarding the components, barriers and facilitators of a successful transition. In addition, some who work in the veteran space have served themselves and so can provide this perspective too.

## 2. Method

### 2.1. Ethical approval

This research received full ethical clearance from the Health Faculties Research Ethics Subcommittee, King’s College London (HR/DP-22/23-33303).

### 2.2. Participants

Stakeholders included individuals who worked at organisations involved in supporting UK service or ex-service personnel (e.g. mental health, financial support, career advice) and policymakers whose work concerned the support available for UK service or ex-service personnel. Stakeholders took part in a series of one-to-one interviews and round-table discussions.

For the individual interviews, individuals/organisations who had exposure to a wide range of ex- servicewomen were identified by both a review of services providing support to ex- servicewomen and through the research team’s professional networks.

To supplement the targeted recruitment of the individual interviews, an open call for participation in two roundtable discussions were advertised on social media and in newsletters that targeted veteran service providers. These were an open call-out inviting anyone who worked with service or ex-service personnel in a support capacity or whose policy work related to the Armed Forces. These roundtables were held in addition to individual interviews to ensure the perspective of a broad range of stakeholders was captured, including ones the research team were not aware of. All interested parties who met the inclusion criteria below were invited to participate (10 people per focus group).

#### 2.2.1. Inclusion criteria

To be eligible to take part in these discussions, individuals needed to be employed in veteran support service; either a third-sector organisation that provided any form of support or advice to ex-service personnel or the National Health Service. Alternatively, they needed to work for the government conducting policy work that pertained to the UK Armed Forces.

### 2.3. Procedure

BC conducted individual interviews on Microsoft Teams, and audio was recorded with consent (video was not captured).

BC familiarised herself with the audio recordings and made comprehensive notes on each interview, key quotes were transcribed verbatim.

LR, BC, and AS hosted two roundtable discussions with additional stakeholders (February and May 2023) online via Microsoft Teams. They lasted two hours with a short break. The stakeholder roundtable discussions were audio recorded with consent, and comprehensive notes were made by all members of the research team, which were then merged into one master document.

### 2.4. Materials

A semi-structured interview guide (Table 1) for the individual interviews was co-developed by the research team and a Patient and Public Involvement and Engagement (PPIE) group comprised of five ex-servicewomen and led by VL, who has also served in the UK Armed Forces. The questions covered the following topics: components of a successful transition to civilian life, including the barriers and facilitators to a successful transition, ex-servicewomen support needs, gender differences and reflections on the current support provision.

**Table 1.**
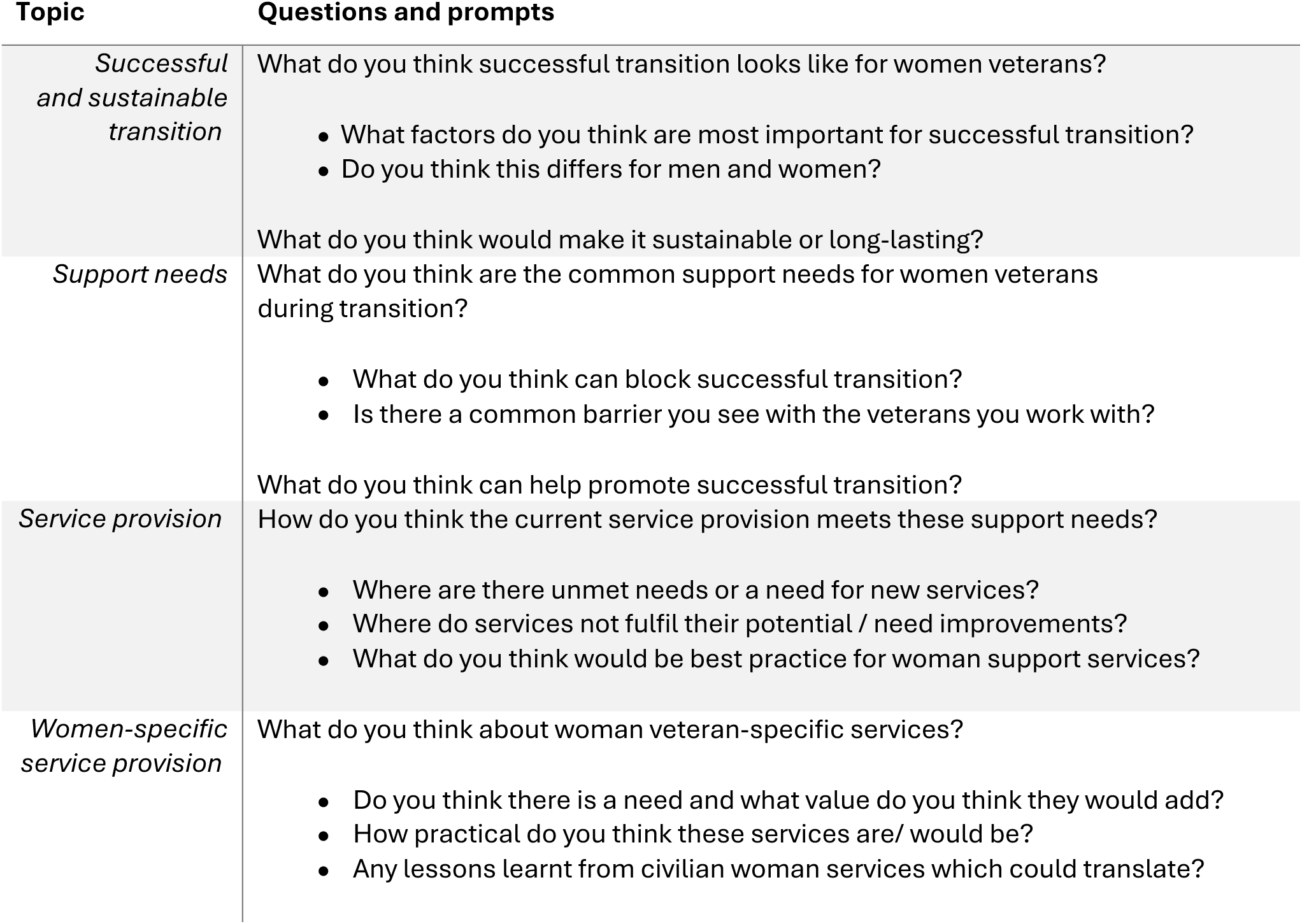
Interview guide for stakeholder discussions.

For the roundtable discussions, the same broad questions from the interview guide were used, and prompts were utilised when needed.

If the stakeholders had served in the UK Armed Forces themselves, they were encouraged to discuss their personal experiences in the roundtable or interviews in addition to their professional perspectives if they felt comfortable doing so.

### 2.5. Analysis

The stakeholder discussions were analysed using a type of thematic analysis known as framework analysis (17). This involved a five-step process of familiarisation with the data, identifying a thematic framework through the use of codes, mapping the study data against these codes, charting to summarise the codes, and interpreting themes from the codes.

BC and LR developed the list of codes and co-produced themes, checking for agreement at the different stages. If there was disagreement, this was resolved with further discussion and if necessary, it was raised with the wider team. These themes were presented to the wider research team and to the PPIE group. The themes were refined based on their feedback.

## 3. Results

### 3.1. Demographic Information

In total, 28 individuals from 22 different organisations participated in this study. For a list of organisations that participated, please see Table 2. Half of the stakeholders (n=14) disclosed that they had served or were currently serving in the UK Armed Forces. There were 23 women and 5 men.

**Table 2.**
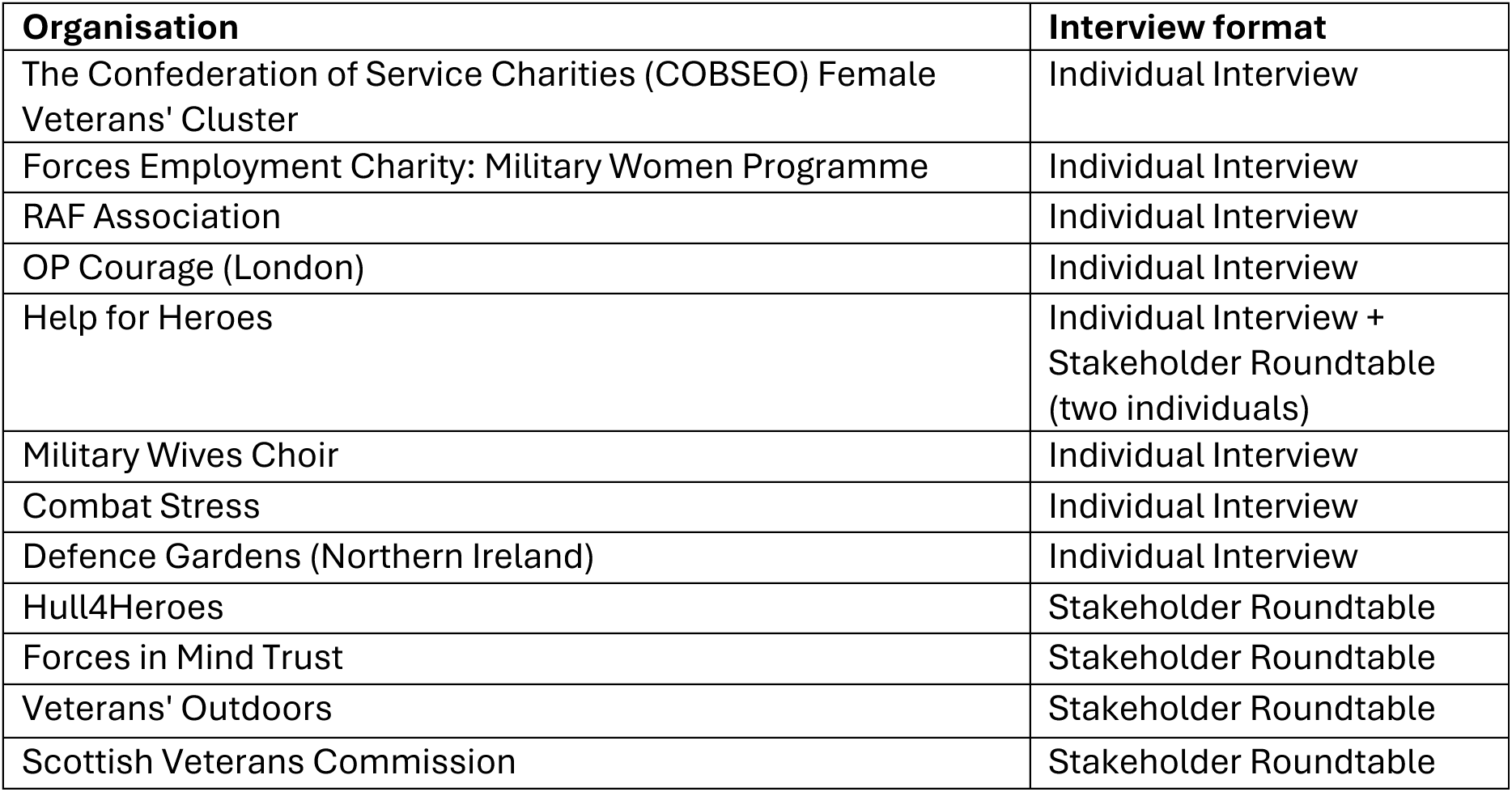
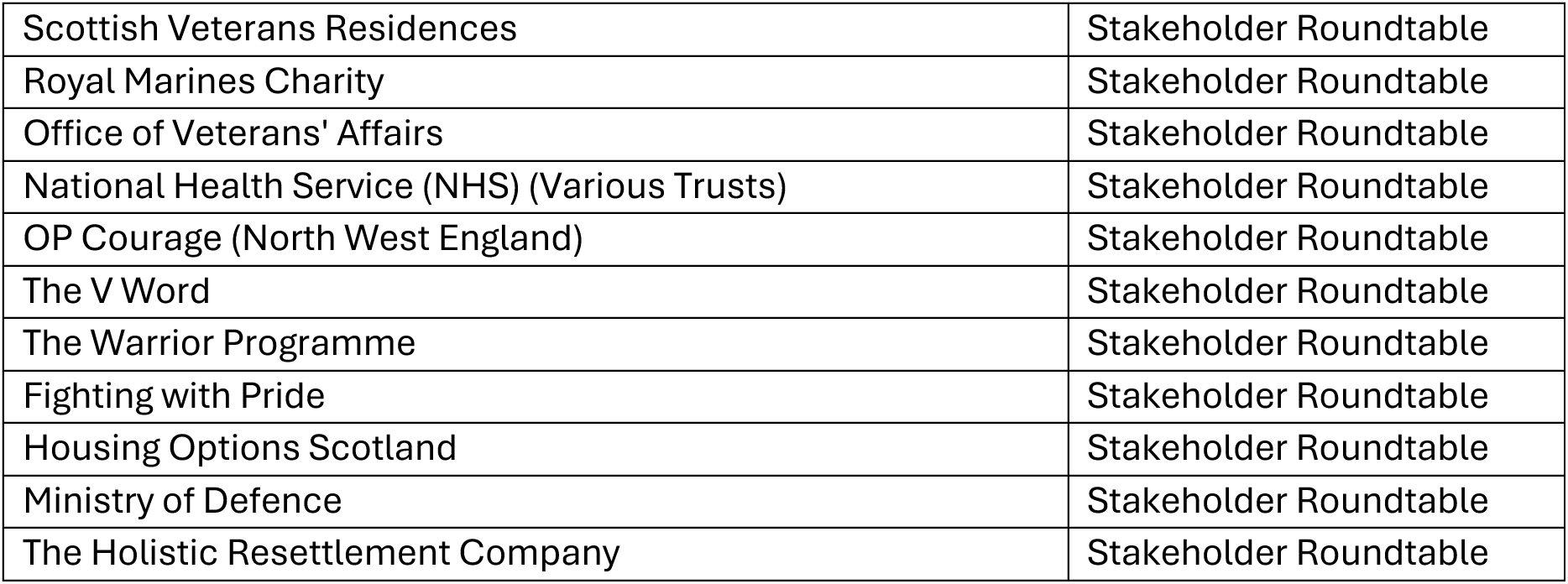
The organisational affiliation of the stakeholders who took part in either an individual interview or a roundtable discussion.

| Organisation | Interview format |
| --- | --- |
| The Confederation of Service Charities (COBSEO) Female Veterans' Cluster | Individual Interview |
| Forces Employment Charity: Military Women Programme | Individual Interview |
| RAF Association | Individual Interview |
| OP Courage (London) | Individual Interview |
| Help for Heroes | Individual Interview + Stakeholder Roundtable (two individuals) |
| Military Wives Choir | Individual Interview |
| Combat Stress | Individual Interview |
| Defence Gardens (Northern Ireland) | Individual Interview |
| Hull4Heroes | Stakeholder Roundtable |
| Forces in Mind Trust | Stakeholder Roundtable |
| Veterans' Outdoors | Stakeholder Roundtable |
| Scottish Veterans Commission | Stakeholder Roundtable |
| Scottish Veterans Residences | Stakeholder Roundtable |
| Royal Marines Charity | Stakeholder Roundtable |
| Office of Veterans' Affairs | Stakeholder Roundtable |
| National Health Service (NHS) (Various Trusts) | Stakeholder Roundtable |
| OP Courage (North West England) | Stakeholder Roundtable |
| The V Word | Stakeholder Roundtable |
| The Warrior Programme | Stakeholder Roundtable |
| Fighting with Pride | Stakeholder Roundtable |
| Housing Options Scotland | Stakeholder Roundtable |
| Ministry of Defence | Stakeholder Roundtable |
| The Holistic Resettlement Company | Stakeholder Roundtable |

The interviews lasted between 32 and 53 minutes (the average length was 38 minutes).

### 3.2. Interview Themes

Four overarching themes were identified: ’The conflicting identities of servicewomen’, ’Sexism: women don’t belong in service’, ’The needs of servicewomen’ and ’Successful transition is individualised and all-encompassing’. The last describes the transition process, and the remaining themes outline common challenges faced by ex-servicewomen and their support needs, which influence the transition process.

Figure 1 presents all themes and sub-themes, which are further expanded upon with quotes from the interview transcripts. Key elements of successful transition or barriers or facilitators are highlighted in bold throughout.

**Figure 1.**
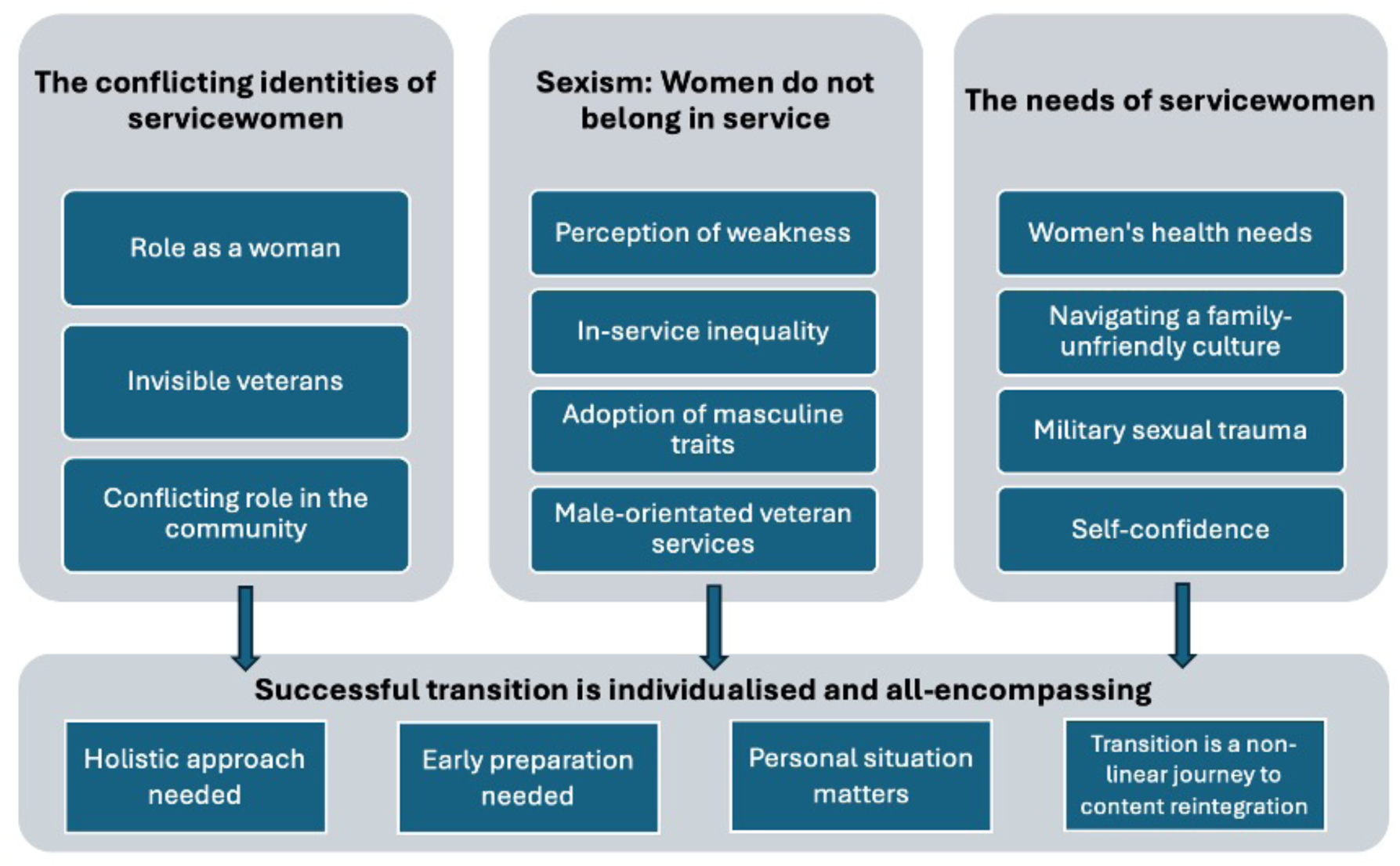
Interview themes and sub-themes

#### 3.2.1. The conflicting identities of servicewomen Role as a woman

Stakeholders described the many roles that a woman took in her lifetime such as a daughter, wife and mother. Those who served in the military then adopted an additional role of ’soldier’ often in conflict with their other responsibilities during and after service. The ’soldier’ role was then diminished as other roles were prioritised. For instance, after service, the ‘daughter’ role may come to the forefront again for those who need to care for elderly parents. Some also become a wife/mother which meant they became a carer in the first instance and some women struggled to maintain a ***sense of their own identity*** beyond a caring role. Stakeholders believed that the unbalanced societal expectations placed on women rather than men to care for their children and to return to their hometown and care for elderly parents, in addition to the stereotype that all veterans are male, as detailed in the next sub-theme, further exacerbated the invisibility of service highlighted in the next sub-theme ’invisible role as a soldier’.

This tendency to be the ’supporter’ meant women felt that their own support needs were secondary, instead prioritising the needs of others first. Women were, therefore, less inclined to ***identify their own support needs*** and seek help for themselves during their transition.

*“And I think there is something unique about woman veterans who are married to other veterans. And I would say the spouses, the spouse veteran, their mental health needs tend to come first. When woman veterans that were married to civilians, it’s been much more kind of, well, you know, she’s been to war.”* – **Stakeholder F.**

##### Invisible role as a soldier

Women’s role as soldiers was often seen as invisible—to the ex-servicewomen herself, to others who served or had served, and to the wider public.

Stakeholders spoke about how typically ex-servicemen strongly associated with being a veteran, which meant they were more likely to maintain their in-service friendships and engage with veteran communities and support services. However, some ex-servicewomen then had to navigate the stereotype that all veterans were men. Stakeholders discussed accounts they had heard or witnessed where women’s service was often doubted. Examples included a woman being told they were wearing their ’husband’s medals on the wrong side’ in reference to their own medals, or a woman who called to claim her war pension, and the adviser assumed that they were a war widow, not an ex-service person.

This disconnect from veteran status also appeared to be internalised, with stakeholders commenting on how ex-servicewomen did not identify as veterans themselves: “*Women are reluctant to identify as veterans and don’t want to engage in veterans’ charities because they don’t perceive themselves as veterans.”* – **Stakeholder D.**

Stakeholders stressed that this led to women being less likely to ***engage in services*** which were veteran-focused. In addition, they reported that ex-servicewomen struggled to identify as a civilian and felt that civilians did not understand them, potentially affected by their adoption of ‘masculine traits’ discussed later. Stakeholders noted that although men too felt this way towards civilian services, women did not identify as either a veteran or civilian, and so these women were left in a state of limbo and subsequently might be less likely to engage in services which were veteran-focused but also feel excluded from civilian services.

##### Conflicting community placement

All stakeholders referenced ***community integration*** as a key element of a successful transition for all ex-service personnel. This refers to social networks as described in the sub-theme ‘Transition is a non-linear journey to content reintegration’ and connections with civilian services. There were mixed views on the differences between ex-servicemen and women in their ability and success in finding a new community after service. Some stakeholders reported that ex-servicewomen struggled to find a community that they could relate to after service, partly due to the difficulty of identifying as a veteran but also their belief that civilians would not understand them.

Conversely, other stakeholders commented that it might be easier for women to transition and integrate into a community if they had children during service. They explained that military parents might have had more contact with civilian services than usual. For instance, prenatal care is facilitated by the NHS, and children attend a civilian school. Therefore, these civilian services and systems were more familiar to these individuals, and they might have been more comfortable using civilian services themselves after service. These stakeholders proposed this as a possible reason why ex-servicewomen reported being less likely to use veteran services.

In addition, stakeholders commented that women were more likely to build social networks and stay in touch with individuals, which helped maintain that sense of community:

*“And I think women are more joiners than men, like in clubs and things like that or…I just think women are a bit more resilient in life, because they have to be, for various reasons.”* – **Stakeholder D**

#### 3.2.3. Sexism: Women don’t belong in service

##### Perception of weakness

Stakeholders described numerous reports of sexism and its potential impact on women accessing support. in particular, they reported male colleagues telling women that they were not able to do the job to the same level as men. Subsequently, they suggested that women felt the need to prove themselves and portray a form of socially acceptable strength. Often, this discouraged them from ***seeking help***, especially for mental health support, as women were reluctant to be seen as the ’weak woman’ making a fuss.

This feeling and subsequent reluctance to ***seek help*** appeared to permeate through into post-service transition, with women still hesitating to seek help to not to appear as ’weak’.

**Stakeholder G**, speaking from personal experience, described how this impacted their help-seeking journey:

*“When I’d get a new posting, I’d know that when they found out I was a woman that was going to that particular job, they’d expect me to be rubbish, they’d expect me to be no good at [physical training] they’d expect me to be terrible at my job, probably think that I was going to sleep with some of the soldiers and that was just the expectation they had so for me to go and for me to even get myself to a level with the guys I had to work ten times hard so it breeds this imposter syndrome because you’re being told all the time you’re not good enough….and that can flow into when you do leave.”*

##### In-service inequality

In addition to the sexist attitudes experienced by women, stakeholders gave examples of the inequality women faced during service including insufficient facilities, poorly designed uniforms, and lack of provision for women-specific issues such as pregnancy and menopause. Although stakeholders could not verify such statements, they reported hearing stories about bullet-proof vests: *“A veteran was telling us that it was only recently that the military created [bullet proof] vests for women. Before that, the women used to have to have breast reductions.”* **-** Stakeholder H

These difficulties faced by women in service further increased the sentiment that women did not belong in the military. This feeling permeated into post-service life, detailed in the sub- theme ’male-orientated veteran services’, which stakeholders felt could be a barrier to women ***seeking help*** from military-affiliated organisations.

##### Adoption of masculine traits

Stakeholders often described the need servicewomen felt to demonstrate more hegemonic masculine traits such as aggression and strength to fit in with the military culture. Similarly, they would partake in ’banter’ to diminish the prejudice that they were weak or unable to do their job. However, when women left and joined a civilian workplace, such learned behaviour risked conflict as women were disliked or viewed negatively due to these masculine traits.

*“Women are a minority group, but hyper-visible. When you come out [of the Armed Forces], you are invisible. What people forget is you have to adapt back again and become more feminine again. People will take your manner and stance a different way – particularly in the workplace… And when you leave you have to change back again and may potentially have to adapt masculinity/femininity…”* – **Stakeholder I**

This further reinforced the perception that women did not completely belong in military or civilian environments, having to adapt to fit a masculine environment in the military but not demonstrating enough femininity for the civilian workplace. This notion of belonging and traditionally gendered traits and behaviour were thought to have a negative impact on transition, particularly in employment, as it created difficulties in ***interpersonal relationships in the civilian workplace*.**

##### Male-orientated veteran services

The sense of women not belonging prevailed into post-service with stakeholders describing the service provision as ’male-orientated’. Many believed most of the services depicted men in their advertisements and highlighted that language such as ’hero’ and ’warrior’ could be perceived as masculine and not inviting to women. On the other hand, services aimed at women were ’overly feminine’, such as the use of the colour pink.

*“Services are male-oriented; I don’t feel like a warrior; I don’t feel like a hunter. Some of the platforms are not aimed at woman veterans, doesn’t resonate and feel it doesn’t apply. Think about the wording, that you use and colours so its inclusive for everyone.”* – **Stakeholder A.**

In addition to the outward appearance and engagement of these services, there was the impression that many services had few women attend or engage with them, which may have been off-putting to women, especially if they had experienced Military Sexual Trauma (MST):

*“I got feedback from one of my woman veteran clients who said: I felt very uncomfortable with some of the comments that the [male] peer workers had made”* – **Stakeholder J.**

Stakeholders had the general impression that these services were often run by men who had served and were of a typical demographic: high rank, white and older. They felt that this created services that were unappealing to women who might feel they do not belong, once again. They also believed these individuals had little awareness or ***empathy for the issues women faced*** and believed this is why some third-sector organisations were reluctant to create women-only services or address issues such as MST.

*“If you walk into [charity name redacted], it’s run by a generation that culturally does not understand what women’s needs are.”* – **Stakeholder K.**

3.2.2. *The needs of servicewomen*

##### Women’s health needs

Stakeholders indicated that ex-servicewomen had specific ***health*** needs, common to all women, both during service and post-service, which could influence how successfully they transitioned out of the UK Armed Forces, including pregnancy, endometriosis, menopause and musculoskeletal difficulties. However, elements of service life and culture exacerbated these health problems. For example, stakeholders pointed out that whilst musculoskeletal problems were experienced by men and women, the inequality in uniform design and equipment exacerbated these issues for women. Until recently (2023), military uniforms were still designed with men in mind, and so the normal features to mitigate the burden of heavy weight from a rucksack or bulletproof vest did not consider the female anatomy. The lack of appropriate uniforms also reiterated the theme: ’women don’t belong’.

*“It’s the body armour, you know, carrying weight and things like that. It impacts women differently. I mean, don’t get me wrong, my knees are just as shot as any other veteran. But, you know, if you have to carry weight, it impacts on our hips.”* – **Stakeholder L.**

For obstetric and gynaecological issues, there was a consensus that whilst the military was trying to accommodate these needs, there was still a poor understanding of how these issues impacted mental health. In addition, there was inadequate provision in facilities to accommodate such issues. For example, stakeholders reported no private areas for servicewomen to pump breast milk.

*“One person I know had really advanced endometriosis…they were still serving had the full hysterectomy actually, had the time off work, was quite well supported in terms of time off but then there was no acknowledgement of well actually now you’re going to have to go on HRT [Hormone Replacement Therapy]… there are all those bits that go on that you know you’re just expected to pick up and be the soldier again…I think within the military there needs to be that education piece because in doing so you can then attract more women through the door.”* - **Stakeholder L.**

The inequality in service with uniform design and the ***lack of understanding*** from military healthcare professionals regarding gynaecological issues meant women sometimes had poor health when they left the military, which impacted their success in civilian life. Stakeholders reported that in extreme cases, musculoskeletal problems can lead to disability and economic inactivity.

##### Navigating a family-unfriendly culture

There was the perception amongst stakeholders that society was changing in regard to the division of labour between men and women when it comes to childcare. However, they felt that in heterosexual partnerships in the military, women were more likely to take on a larger share of the childcare. Despite shared parental leave policies, stakeholders believed it still was not seen as acceptable for the man in a heterosexual relationship to take longer than the statutory two weeks of paternity leave which meant that the prevailing expectation was that the servicewoman would take the longer period of leave. They referenced the pregnancy ban, which meant until 1990, if a servicewoman was pregnant, then she would be discharged from the UK military. They believed whilst this policy was no longer in place, it contributed to these ***childcare expectations***. Starting a family and childcare responsibilities were also cited as the most common reasons stakeholders believed women left the UK Armed Forces. Common reasons included not wanting to be sent away on deployments with young children, the inflexibility of the military to accommodate school pick-ups and childcare in general, and a lack of understanding about post-natal mental health. Whilst these issues are barriers to any service person with children, stakeholders believed women were more likely to be negatively impacted by this due to the incompatibility between the military and child-rearing generated by archaic attitudes of motherhood, which permeated through the military organisation. For those women who felt they had to leave when they had children, stakeholders theorised that ***this lack of choice and autonomy*** over leaving could impede the ***emotional adjustment to transition*.**

*“It’s really challenging to raise children in the military… I was always going away on operations, I had two very young children. I was made to feel after my second child…that I should come back from my maternity leave early…my unit made me feel so bad about being off.”* – **Stakeholder D** talking about their personal experience.

##### Military Sexual Trauma (MST)

There was recognition amongst stakeholders that ***MST***, including sexual harassment and assault, was not uncommon in the UK military and they had some experience of working with ex-servicewomen who had experienced this. MST was identified as a support need that required specialist, ***trauma-informed care*** and had serious implications for women’s mental health.

When asked about their views of women-only veteran services, there were mixed opinions on whether they were warranted but consistently, MST was named as the exception. Stakeholders all agreed that if a woman had experienced MST, they could benefit from women-only veteran services to feel safe. This included having female peer workers and therapists and women-only housing. In reference to a woman-only group they facilitated, one stakeholder explained why they provided this service: *“One of the reasons for the [women-only] group is because of certain issues that people have had before working alongside males.”* – **Stakeholder M.**

##### Self-confidence

***Finding employment*** after leaving the UK Armed Forces was viewed by stakeholders as a common need for all ex-service personnel, irrespective of gender, and often crucial to successful transition. However, stakeholders highlighted that the reasons ex-service personnel sometimes struggled to gain employment after service varied between men and women.

Common difficulties for men included not knowing how to translate military skills into language that civilians would understand for their CV and not knowing how to approach the civilian job market. For women, stakeholders believed it was less about communicating their skills and more about believing they had skills which would be valuable in the civilian world. Stakeholders described a ***lack of self-confidence*** and belief which they felt was more common in women **Stakeholder L** explained, “ They have self-esteem issues…they do not see what a potential employer can see in them.”

In the sub-theme’ perception of weakness’, stakeholders described how women were sometimes viewed by male colleagues as ’less-worthy’ or ’less able’ to do the job, and so women felt they had to work harder to prove themselves. It is possible that this contributed to a lack of self-confidence in civilian life.

#### 3.2.3. Successful transition is individualised and all-encompassing

##### Holistic approach needed

When asked about their views of the current service provision, many stakeholders commended the wealth of services available to ex-service personnel. However, they noted that there appeared to be a gap for a ’holistic’ approach to transition, meaning there was no service which offered a person-centred approach (i.e. which covered all needs ranging from practical support such as housing to emotional counselling for adjusting to civilian life). They referenced the Career Transition Partnership (CTP) (Ministry of Defence’s former provider (until March 2024) of Armed Forces Resettlement), which provides support with finding employment after service and noted that it might prepare you to write a CV and other organisations may help you find a home, but participants suggested that few, if any, prepared an individual for the ***emotional side of leaving the UK Armed Forces***. They further explained that leaving the Armed Forces involved the loss of friends, a certain way of life and purpose for both men and women. One stakeholder likened it to a grieving process: *“The human factor is missing, understand the emotional and human. Leaving the military is going through the grief cycle. Even if you are excited to move on, even with a personal choice, it’s a grief cycle, this is even worse for people who don’t choose, for example medical discharge.”* - **Stakeholder A**

##### Personal situation matters

Repeatedly, stakeholders emphasised that successful transition for all ex-service personnel was individual and dependent on what that person needed and their personal situation.

*“A successful transition is one that is measured (i.e. not done in a hurry), one that takes advantage of all the resettlement services that exist and one that gives the individual the transition that they need, not the transition that the service necessarily template for them so it takes into account the variables for that individual because individuals will have very different needs depending on the circumstances that they’re going into civilian life in.”* – **Stakeholder B**

There were several factors that stakeholders anecdotally listed, which they believed made the transition more difficult for women. This included having a partner who was still in service, which they believed could be problematic as women became the ’military wife’, demoting women’s own service experiences, as mentioned in the sub-theme, ’role as a woman’.

Stakeholders explained further that military spouses often lived on base even after their own service had ended, leading to confusion around their identity as women tried to adapt to a new civilian identity, but this was a juxtaposition to their environment and the local community. The reason for discharge was viewed as also being pertinent to transition and an individual’s personal situation. For those (men and women) who were medically discharged, stakeholders explained that this was often sudden and unwanted, so coming to terms with this, in addition to a potentially permanent injury, could be detrimental to a person’s mental health.

##### Transition is a non-linear journey to content reintegration

Stakeholders agreed that the time taken for someone to ’successfully transition’ out of the UK Armed Forces was individual, with some leavers achieving this in the first two years and others never fully transitioning:

*“Transition starts at different stages for different people depending on their reasons for leaving and can last for as long as it takes for an individual to genuinely say ’I’m **content** in myself’.”* - **Stakeholder C.**

All stakeholders concurred that ***reintegration into society*** was a key component of a successful transition. A few stakeholders, including those who had served themselves, noted that some ex-service personnel engaged in a veteran service or charity for a prolonged period either as an employee or beneficiary, meaning their support network remained predominantly ex-military. Stakeholders believed this could be a barrier to fully reintegrating into the civilian world as they never fully emotionally accepted, they had left the Armed Forces. Stakeholders believed women were less likely to do this than men, for reasons noted in sub-theme ‘male- orientated veteran services’, and so this facilitated a better integration into civilian life, mainly due to the development of civilian social networks.

Some stakeholders, however, noted that the process of reintegrating into society was not linear, with several steps backwards, such as changing career paths multiple times before a successful transition is achieved. More than half of the stakeholders felt that the lack of linearity was more than often the case for women compared to men. Some provided the example of women leaving the Armed Forces to have and care for children and stated that women may then not prioritise employment. In such situations, women did not engage with CTP and career advice and support were needed much later when the military no longer provided it.

##### Early preparation needed

A recurrent theme amongst stakeholders was the need for all serving personnel to ***prepare early*** for eventually leaving the military. Although resettlement opportunities are available for up to two years after an individual decides to leave the military, stakeholders believed this should not be the only preparation that is done. Stakeholders explained that a person should be thinking and preparing about leaving when they join. For example, making the most use out of the military’s education and training, and trying to integrate into the civilian community whilst in service. This meant when individuals did leave, the emotional side of transition, as mentioned in the sub-theme ’holistic approach needed’, was not as difficult. Stakeholders reported this would also be beneficial to individuals who were medically discharged and did not expect to be leaving service.

## 4. Discussion

This research aimed to understand how stakeholders working with veterans defined a successful transition out of the UK Armed Forces into civilian life. No singular definition emerged as to what constituted a successful transition. However, key components of a successful transition were highlighted by stakeholders which formed steps towards one definition. In summary, a successful transition is one where early preparation enables individuals to consider all elements of transition, including the emotional side, in order to become content with themselves and re-integrate into civilian life. What was more prominent in the findings was stakeholders’ reports of barriers to a successful transition. This is perhaps unsurprising given that stakeholders see women in a support capacity and hence will engage with female veterans experiencing difficulties. Nonetheless identifying female needs and barriers experienced can still provide a critical insight, in particular examining where barriers arise and offering potential solutions or interventions. Additionally, whilst this study focused on transition, many of the barriers were derived from in-service experience. Stakeholders agreed that the in-service experience of women differed from those of their male counterparts, which affected their outcomes post-service.

### 4.1. Results in relation to previous literature

Sexism and self-confidence: The perception of being weak and not fit for the job whilst in service was an experience stakeholders believed was particularly common for their clients who were women. This echoes evidence from a survey of UK ex-servicewomen, with 40.1% reporting they felt the need to be better than male colleagues to get the same recognition, and 19.9% felt they were treated differently which led to a lack of self-confidence (11). This experience has also been noted in other traditionally male-dominated industries such as the UK fire service where women firefighters reported feeling subordinate and that their work and abilities were seen as less valuable than their male counterparts (18). This suggests it is not a problem unique to the military but is possibly attributable to a male-dominated environment. This in-service experience impacted the needs of women when they left. Stakeholders believed women, more than men, had low self-confidence when applying for civilian jobs, possibly attributable to these perceptions from colleagues, and this meant employment support offered by veteran services may benefit from tailoring its approach differently across ex-servicemen and women.

Physical health needs of women: Stakeholders explained that women’s specific health needs, such as menopause and endometriosis, were still poorly understood by the military. In a survey of UK servicewomen exploring perimenopause, 54% reported that access to a suitable healthcare professional was difficult and 35.7% reported not being ’heard’ when they had spoken to a healthcare professional about their symptoms (19). Through analysis of free-text responses, participants reported that they often felt male colleagues and healthcare professionals, more so than women, lacked understanding of the psychological and physical effects of perimenopause which impacted their occupational performance. This is perhaps unsurprising given that in the general population, there is limited awareness of some women’s health issues. In a qualitative study, patients in the general population with endometriosis described experiences of being told by doctors that the pain was psychosomatic; the women in the study believed there was a systemic lack of awareness of the condition, but they also felt that pain experience was gendered, and women’s pain was often dismissed (20). Since the military requires a healthy workforce to deploy, failing to provide healthcare for women properly will have an impact on the military’s ability to carry out duties vital to national security.

Similarly, there should be more awareness of these conditions amongst service providers to better support ex-servicewomen with these health conditions to improve their transition experience and quality of life.

Potential benefit of conflicting roles: Although barriers existed, rooted in misogyny and inequality, stakeholders described ex-servicewomen as being generally resilient and resourceful. The various identities and roles women have over their life course and military service appeared to facilitate an easier adaptation to civilian life compared to men. For instance, a knowledge of civilian services like schools and the NHS from having children meant a level of familiarity with these services. Similarly, stakeholders believed women were more able to form social networks after leaving than men, which had a positive impact on wellbeing. Previous research has suggested that transition is harder for ex-service personnel who have a more salient military identity (6) and so having multiple other identities may be protective for ex- servicewomen. Similarly, although women do not access veteran services as much as men (13), on the whole, they seek formal help more than men (21) but access civilian services instead. This could be a result of those services not feeling inclusive to women, their familiarity with civilian services seems to be protective in that they still feel able to access support if they need to, which is important to acknowledge. Although it is reassuring that women are still using civilian services, they are potentially missing out on the benefits that veteran services can provide such as quicker waiting lists and unique military knowledge, meaning women are potentially still at a disadvantage to their male peers who do utilise these associated benefits.

Reason for leaving: Stakeholders believed that for all veterans, being medically discharged was often more difficult mentally than leaving by choice because of the lack of control and in some cases, life-changing injuries. This is supported by evidence which showed that PTSD and CMD were associated with being medically discharged and ‘other unplanned leaving’ in UK ex-service personnel (22), although it is possible that some of these individuals were medically discharged for mental health in the first place. This is particularly pertinent to women who are more likely to leave the UK military for medical reasons (23). The lack of control and autonomy and the impact on wellbeing is also important to consider for those women who feel the military culture and lack of flexibility may make having children in-service untenable, forcing them to leave the Armed Forces.

### 4.2. Strengths and limitations

The value in talking to stakeholders is that they can provide a broad overview of ex- servicewomen’s experience as they have worked with a wide range of women (different ranks, service branches and circumstances). Stakeholders reported several anecdotes from the ex- servicewomen they had worked with. However, it is not possible to know which era these experiences refer to and whether they are reflective of current military culture and policy, for example, the removal of automatic dismissal in the case of pregnancy in 1990 (24) and the new zero-tolerance policy on sexual offences and sexual relations between trainees and instructors in 2022 (25). Nonetheless, it was apparent that poor historic experiences could impact women’s civilian lives and still requiring support. There may also be a bias in the experiences recounted, as many stakeholders worked in support services, they might see a select group of women who have had a poor experience in service such as harassment or discrimination or had difficulty when they left (such as needing housing, employment) which led them to seek support. In addition, some of these stakeholders had served themselves so their own personal experiences transpired in the interviews. Nevertheless, the stakeholders involved in this study were from a variety of organisations that covered several sectors and all four regions of the UK, which may help overcome some of the selection biases that can occur when recruiting ex- servicewomen themselves.

### 4.3. Implications for policy, practice and future research

The experiences of harassment, discrimination and MST faced by servicewomen contribute to a feeling of not belonging, which permeates into civilian life, affecting their transition and sometimes eclipsing the positive memories of their service. These experiences are arguably exacerbated by a culture that encourages hegemonic masculinity. Military culture is entrenched in centuries of historical context and shaped around the needs of war, and as such, military training arguably emphasises traits such as emotional stoicism, dominance, violence and heterosexual desire (26). This male-dominating culture still persists today and was criticised in The Atherton Report, which was the first inquiry into the experiences of “women in the armed forces from recruitment to civilian life” (2021)(27). There is a potential link between such characteristics and the discrimination and harassment of women. The UK Ministry of Defence has set out policies and measures to tackle sexual offending and has a zero-tolerance stance to such behaviour (28). These policies focus on identifying the root cause of offending, identifying at-risk individuals (both survivors and perpetrators), and ensuring people feel comfortable calling out unacceptable behaviour. As such, policies are critical and fundamental to creating an inclusive and diverse workforce. However, we argue that to address harassment and discrimination in the military appropriately, attention also needs to consider cultural change. Other behaviours, such as inappropriate humour, as identified in the current study, might go unreported as they are often dismissed as they do not break any laws per se.

Nevertheless, these should not be ignored as they all contribute to the perpetuation of a hegemonic masculine culture and group attitudes towards women. Although this culture has some positive implications such as a supportive ’brotherhood’ (29), such a culture can lead to men minimising or hiding emotion (29) and prevent male veterans from seeking help (26).

Therefore, interventions to address cultural change are paramount in improving the in-service experience for all personnel whilst maintaining an emphasis on strength and robustness, which are intrinsic to the military mindset.

Although direct discrimination from colleagues played a large role in women’s perception that they did not belong, stakeholders believed that indirect discrimination or inequity in the facilities, such as no sanitary bins being available, contributed to this perception. The military has already gone some way in helping women feel a part of the organisation by changing gendered terms; aircraftsman was changed by the Royal Air Force in 2022 to ’air specialist’ (30) and body armour has now been redesigned for women (31). However, the results of this study indicate that similar steps are yet to be taken by veteran services as many stakeholders believed there was a lack of awareness by these services of women’s needs and the marketing was often geared towards men. There are various veteran awareness training programmes that exist, yet we are not aware of any that have a module on the specific needs of women outside of training to support those who have experienced MST. Incorporating a module into training programmes would be a low-cost intervention that could help make women feel more recognised and supported by the veteran services and facilitate a more successful transition.

This could also mitigate the need for women-only veteran services which stakeholders believed was only necessary for very specific issues such as MST where women might need segregated spaces to feel safe and comfortable to express thoughts freely.

To fit into a male-dominated environment, stakeholders described the tendency for servicewomen to adopt more traditional masculine traits. Once they joined a civilian workplace, they felt they were again not accepted as they were viewed as ’harsh’ or ’brash’ and did not conform to a traditional feminine ideal. This perception of women is not exclusive to ex- servicewomen; analysis of British media shows gendered differences in descriptions of men and women in leadership roles; broadcasters focus on appearance and domestic and childcare duties when discussing women and comment on how likeable they are, which was often more negative when they were viewed as masculine (32). However, for ex-servicewomen, this systemic sexism in the workplace might be more challenging because of adaptations they have made to conform to the military culture. Therefore, although cultural change interventions in service could be beneficial as seen in other organisations (33), and this might improve as the number of women serving in the UK Armed Forces increases, the systemic discrimination of women in the workplace cannot be ignored as this, impacts women’s experience of transition, in particular job satisfaction and career prospects.

### 4.4. Conclusions

Despite no singular definition of successful transition being identified, the feedback from stakeholders has provided valuable insight into barriers which may hinder successful transition. Through these, we can identify points of intervention and prevention to foster a more successful transition for women going forward. This study also aimed to understand the experience of transition through the lens of ex-servicewomen, and we can conclude that the dynamics of gender and negative experiences such as misogyny and inequality can overshadow positive aspects of women’s service. Where there are universal experiences or barriers to success, the solutions to these difficulties may require a different approach for men and women. More exploration of the lived experience of ex-servicewomen is needed. However, we can conclude from stakeholder reports that experiences of sexism, misogyny and feelings of estrangement in ex-servicewomen prevail in civilian life through workplace discrimination and exclusionary veteran services. Therefore, interventions should target both in-service discrimination and post-service provision but acknowledge the broader ecological environment where sexism and misogyny still prevails in society, which may need longer-term population-level interventions.

## Data Availability

Data not available due to ethical restrictions.

## Acknowledgements

We are grateful to our participants for contributing their data to this study and for being candid and forthcoming in discussions.

## Copyright

For the purpose of open access, the author has applied a Creative Commons Attribution (CC BY) licence to any Author Accepted Manuscript version arising.

## Footnotes

1 In the UK, veterans or ex-service personnel are defined as anyone who has served for at least one day in His Majesty’s Armed Forces (Regular or Reserve) or Merchant Mariners who have seen duty on legally defined military operations. https://assets.publishing.service.gov.uk/media/5e79ebdad3bf7f52f9ee0b42/6.6409_CO_Armed-Forces_Veterans-Factsheet_v9_web.pdf

